# Tunable, soft robotics-enabled patient-specific hydrodynamic model of aortic stenosis and secondary ventricular remodeling

**DOI:** 10.1101/2022.09.12.22279793

**Authors:** Luca Rosalia, Caglar Ozturk, Debkalpa Goswami, Jean Bonnemain, Sophie X. Wang, Benjamin Bonner, James Weaver, Christopher T. Nguyen, Ellen T. Roche

**Author notes:** Corresponding authors: Christopher T. Nguyen., Ellen T. Roche.

## Abstract

Aortic stenosis (AS) affects approximately 1.5 million people in the US and is associated with a 5-year survival rate of 20% if untreated. In these patients, aortic valve replacement is performed to restore adequate hemodynamics and alleviate symptoms. The development of next-generation prosthetic aortic valves seeks to provide enhanced hemodynamic performance, durability, and long-term safety, emphasizing the need of high-fidelity testing platforms for these devices. We propose a soft robotic model of AS capable of recapitulating patient-specific hemodynamics of AS and secondary ventricular remodeling, validated against clinical data. The model leverages 3D printed replicas of each patient’s cardiac anatomy and patient-specific soft robotic sleeves to recreate the patients’ hemodynamics. An aortic sleeve allows mimicry of AS lesions due to degenerative or congenital disease, while a left ventricular sleeve recapitulates loss of ventricular compliance, and impaired filling associated with AS. Through a combination of echocardiographic and catheterization techniques, this system is shown to recreate clinical metrics of AS with greater controllability compared to methods based on image-guided aortic root reconstruction, and parameters of cardiac function which rigid systems fail to mimic physiologically. Finally, we demonstrate the use of this model for the evaluation of transcatheter aortic valves in a subset of patients with diverse anatomies, etiologies, and disease states. Through the development of a high-fidelity model of AS and secondary remodeling, this work pioneers the use of patient-specific soft robotic platforms of cardiovascular disease, with potential application in device development, procedural planning, and outcome prediction in industrial and clinical settings.

**One Sentence Summary:** A high-fidelity, soft robotics-driven model recreates patient-specific biomechanics and hemodynamics of cardiovascular disease.

## INTRODUCTION

Aortic stenosis (AS) is the narrowing of the aortic valve orifice due to reduced mobility of the valve leaflets. It arises as a result of inflammatory processes akin to those driving atherosclerosis, whereby endothelial damage due to mechanical stress and other biological processes induces fibrosis, thickening, and calcification of the valve leaflets *(1, 2)*. Although AS affects the elderly population disproportionately, its onset and progression can be dramatically accelerated by existing underlying congenital defects – such as bicuspid aortic valve (BAV) disease – which occurs when two aortic valve leaflets are fused together *(3)*. Hemodynamically, the narrowing of the aortic valve orifice gradually leads to elevated transaortic pressure gradients *(4, 5)*. The increased afterload (or pressure overload) results in higher left ventricular (LV) systolic pressures and a reduction in the volume ejected at each heartbeat (stroke volume, SV) leading to drops in cardiac output and the onset of symptoms such as angina and exertional syncope *(6, 7)*. In two thirds of AS patients, pressure overload drives LV remodeling, resulting in loss of LV compliance and eventually in diastolic and systolic dysfunction *(8–10)*. This complication of AS causes higher mortality and rehospitalization rates after aortic valve replacement, and may eventually lead to heart failure *(11–13)*.

AS currently affects approximately 1.5 million people in the US and is associated with a 5-year survival rate of 20% from the onset of symptoms, if untreated *(14, 15)*. To date, there are no effective pharmacological treatments for AS, and it is estimated that 80,000-85,000 aortic valve replacement procedures are performed every year in the USA *(16, 17)*. The prosthetic aortic valve market (valued at $6.9 billion in 2021) is rapidly expanding and is projected to reach $19.7 billion by 2031 *(18)*. Next-generation prosthetic aortic valves are currently under development, aiming to enhance hemodynamic performance, durability, and long-term safety *(19)*, which emphasizes the need for high-fidelity patient-specific platforms for the evaluation of these devices *(20, 21)*. Unfortunately, the majority of hydrodynamic models currently used for functional evaluation of prosthetic valves rely on rigid, idealized components and fail to recreate patient-specific anatomies and hemodynamics *(22)*.

Recently, hydrodynamic platforms that integrate patient-specific aortic replicas have been developed for studies of congenital heart disease *(23)*, aortic dissection *(24, 25)*, and AS *(26, 27)*. Specifically, Kovarovic *et al*. proposed a patient-specific model that integrates molded replicas of patient-specific aortic root and calcific valve geometries obtained from computed tomography (CT) data with a rigid pulse duplicator system *(27)*. Similarly, Haghiashtiani *et al*. combined image-guided aortic root models with a rigid pulsatile pumping system *(28)*. In their work, they leverage the multi-material 3D-printing (MM3DP) approach first demonstrated by Hosny *et al*. to manufacture anatomical models of calcified valves, with the advantage of enhanced prototype versatility compared to mold-based techniques *(29)*. Nevertheless, these models are largely dependent upon the quality of the patients’ CT images: low spatiotemporal-resolution CT scans and inaccurate image segmentation would greatly compromise the accuracy of these models and their ability to reliably recapitulate patient-specific hemodynamics. Due to the lack of tunability, several design-manufacturing-testing iterations could be required to obtain a high-fidelity system, hindering the translatability of these models to clinical or industrial settings. Furthermore, by relying on traditional pumps or pulse duplicators, these systems are unable to model changes in the LV compliance caused by remodeling secondary to AS, which is observed in most of these patients, severely limiting the clinical relevance of these models.

Leveraging our previous work, in which we demonstrated the ability of a non-patient-specific aortic sleeve to recreate the hemodynamics of AS in a porcine model *(30)*, we propose a soft robotics-enabled 3D printed anatomical hydrodynamic system that is capable of recreating AS and congenital defects in a patient-specific fashion. In addition, using an analogous design workflow, we develop a patient-specific soft robotic LV sleeve that allows us to mimic changes in cardiac function observed in these patients, simulating longitudinal disease progression parametrically. We demonstrate that our soft robotic aortic sleeve can be controlled to recreate patient-specific hemodynamics of AS more accurately than current methods. Moreover, we showcase the ability of our model to mimic a patients’ hemodynamic changes in LV compliance and diastolic function, creating a high-fidelity and clinically relevant model of AS and ventricular remodeling.

This soft robotics-enabled model of both aortic and LV hemodynamics of relevance in AS demonstrates the advantage of increased tunability over more traditional approaches, paving the way towards high-fidelity testing platforms for the evaluation of cardiac devices, personalized device selection, and outcome prediction.

## RESULTS

### Workflow and architecture for soft robotics-enabled model to recreate the anatomy and hemodynamics of patients with AS

We retrospectively selected 13 patients with a diagnosis of AS who had undergone transthoracic (TTE) or transesophageal (TEE) echocardiography, or a combination thereof, as well as CT imaging for hemodynamic and anatomic evaluation. Our patient cohort had a broad spectrum of functional and structural characteristics relevant to AS, as summarized in Table 1. In this study population of 13 patients (6 female; age: 78 ± 13 years; BSA range: 1.67-2.23 m^2^), the aortic annular diameter ranged from 22 to 32 mm. Two of the selected patients had a bicuspid aortic valve (BAV) anatomy, 6 were diagnosed with severe AS, 12 showed some evidence of aortic regurgitation, 8 displayed thickening of the LV wall, and 4 had a left ventricular ejection fraction (LVEF) lower than 40% *(31)*. Further details can be found in table S1.

**Table 1.** Summary of patients’ anatomical and hemodynamic characteristics. F: female. M: male, BSA: body surface area, d: diameter, LVEF: left ventricular ejection fraction. Thresholds for AS and regurgitation severity *(32)* and wall thickness *(8)* from the literature.

| Patient | Sex | Age range | BSA (m<sup>2</sup>) | Annulus d (mm) | Valve anatomy | Severity of stenosis | Regurgitation | Wall thickness | LVEF (%) |
| --- | --- | --- | --- | --- | --- | --- | --- | --- | --- |
| 1 | F | 86-90 | 1.91 | 24 | Tricuspid | Moderate | Trace | Normal | 43.2 |
| 2 | M | 91-95 | 2.05 | 28 | Tricuspid | Severe | Mild | Normal | 34.4 |
| 3 | M | 86-90 | 1.93 | 23 | Tricuspid | Severe | Trace | Increased | 40.6 |
| 4 | F | 86-90 | 1.67 | 22 | Tricuspid | Moderate | Trace | Increased | 73.6 |
| 5 | F | 86-90 | 1.81 | 24 | Tricuspid | Severe | Mild | Increased | 47.2 |
| 6 | M | 76-80 | 2.19 | 25 | Tricuspid | Severe | Trace | Increased | 71.8 |
| 7 | F | 86-90 | 1.89 | 26 | Tricuspid | Moderate | Mild | Increased | 56.2 |
| 8 | M | 76-80 | 2.21 | 23 | Tricuspid | Moderate | Trace | Normal | 51.2 |
| 9 | F | 76-80 | 1.85 | 25 | Tricuspid | Severe | Trace | Normal | 44.0 |
| 10 | M | 76-80 | 2.1 | 27 | Tricuspid | Moderate | No | Increased | 29.5 |
| 11 | F | 86-90 | 1.87 | 22 | Tricuspid | Mild | Trace | Increased | 41.1 |
| 12 | M | 51-55 | 2.23 | 30 | Bicuspid | Severe | Trace | Increased | 32.3 |
| 13 | M | 46-50 | 2.08 | 32 | Bicuspid | Moderate | Trace | Normal | 31.3 |

Figure 1 summarizes the workflow and overall architecture of our model developed based on patient hemodynamics and imaging. Clinical data, including CT, TTE/TEE, and catheterization data, were obtained from AS patients (Fig. 1A). We first segmented the CT images to create 3D anatomic models of patients’ LV and aortas (Fig. 1B, 2A), which we 3D printed with a soft elastomeric photopolymer resin (Fig. 1C, fig. S1). Then, we used CT images to design patient-specific soft robotic LV and aortic sleeves (Fig. 1C, fig. S2). When combined with the patient specific 3D models and our *in vitro* hydrodynamic model (Fig. 1D, fig. S3), the soft robotic LV sleeve provided the contractile force necessary to generate patient-specific systolic pressure and flows as well as modulation of LV compliance seen in the spectrum of pressure overload, while the soft robotic aortic sleeve provided morphologic mimicry and recapitulation of patient specific hemodynamics. Ultimately, this personalized model allowed for testing of hemodynamic changes induced by transcatheter aortic valve replacement (TAVR) under different conditions (Fig. 1D).

**Fig. 1.**
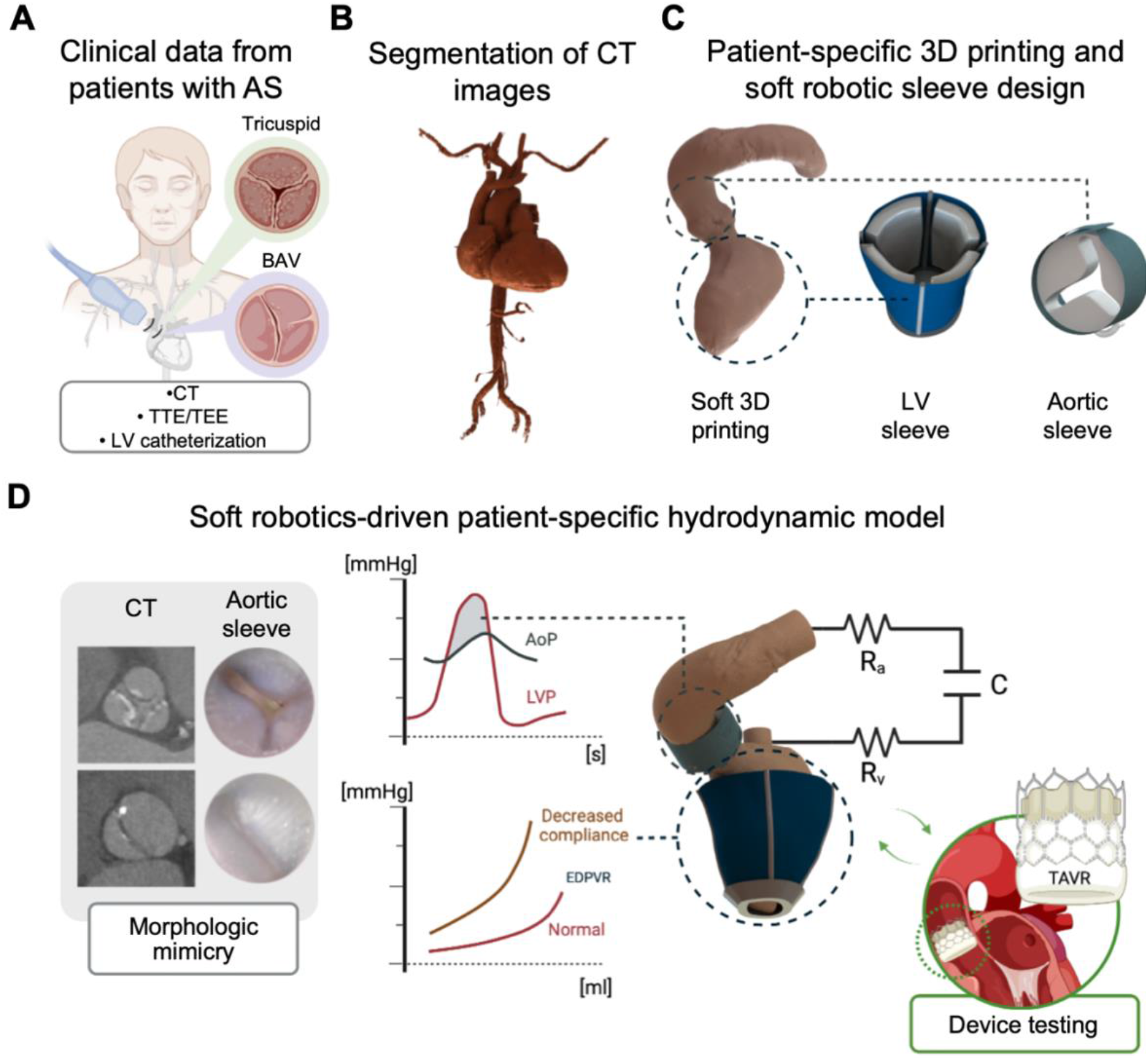
Architecture of the soft robotic patient-specific hemodynamic model of AS and ventricular dysfunction. (**A**) The model is based on clinical data of patients with AS with diverse anatomies and disease etiologies, such as degenerative tricuspid AS and bicuspid aortic valve disease (BAV). These patients underwent CT imaging, echocardiography, and, in some instances, LV catheterization. (**B**) Segmentation of the CT images for the design of 3D anatomical models of each patient. (**C**) Key model elements: soft-material 3D printed LV and aorta anatomical replicas and patient-specific LV and aortic sleeves. (**D**) Schematic of the soft robotics-driven hydrodynamic model. The system is designed to recreate the patients’ hemodynamics and AS morphologies and is ultimately intended as a tool for device evaluation. TTE: transthoracic echocardiography. TEE: transesophageal echocardiography. AoP: aortic pressure. LVP: left ventricular pressure. EDPVR: end-diastolic pressure-volume relationship. R_a_: arterial resistance. R_v_: venous resistance. C: systemic compliance. TAVR: transaortic valve replacement.

The platform designed and developed in this work is ultimately intended for high-fidelity testing and evaluation of medical devices for AS, procedural planning and outcome prediction, as well as product selection tailored to each patient’s anatomy, hemodynamics, and disease state. We demonstrate potential use of our model in this area through implantation of a transcatheter aortic valve replacement (TAVR) valve in a subset of our study cohort.

### Soft robotic aortic sleeve recapitulates patient-specific morphology of AS and congenital valvular defects

The soft robotic aortic sleeve enabled us to recreate the valve lesion morphology of each individual patient with high fidelity. Figure 2A shows a comparison, for each patient, of the aortic valve cine CT images and of the aortic cross-sections of our model under actuation of the aortic sleeve, captured using an endoscopic camera in the system. These images demonstrate that our sleeve can qualitatively recreate a range of patient-specific anatomies, including those of degenerative AS and BAV, with high accuracy. We then superimposed the CT and model images of the orifice area for each patient and calculated the Sørensen–Dice similarity coefficient (DSC) for a quantitative comparison *(33)*. An example of superimposed CT and aortic sleeve images is illustrated in Fig. 2B, where the white area indicates the region of perfect overlap between the two images, while the cyan or red areas indicate regions of image mismatch. All images used for calculation of DSC score can be found in fig. S4. The average DSC across the 13 patients modeled was 0.88 ± 0.05 (Fig. 2C), indicating excellent overlap between native valve morphology seen on CT and our personalized aortic sleeve.

**Fig. 2.**
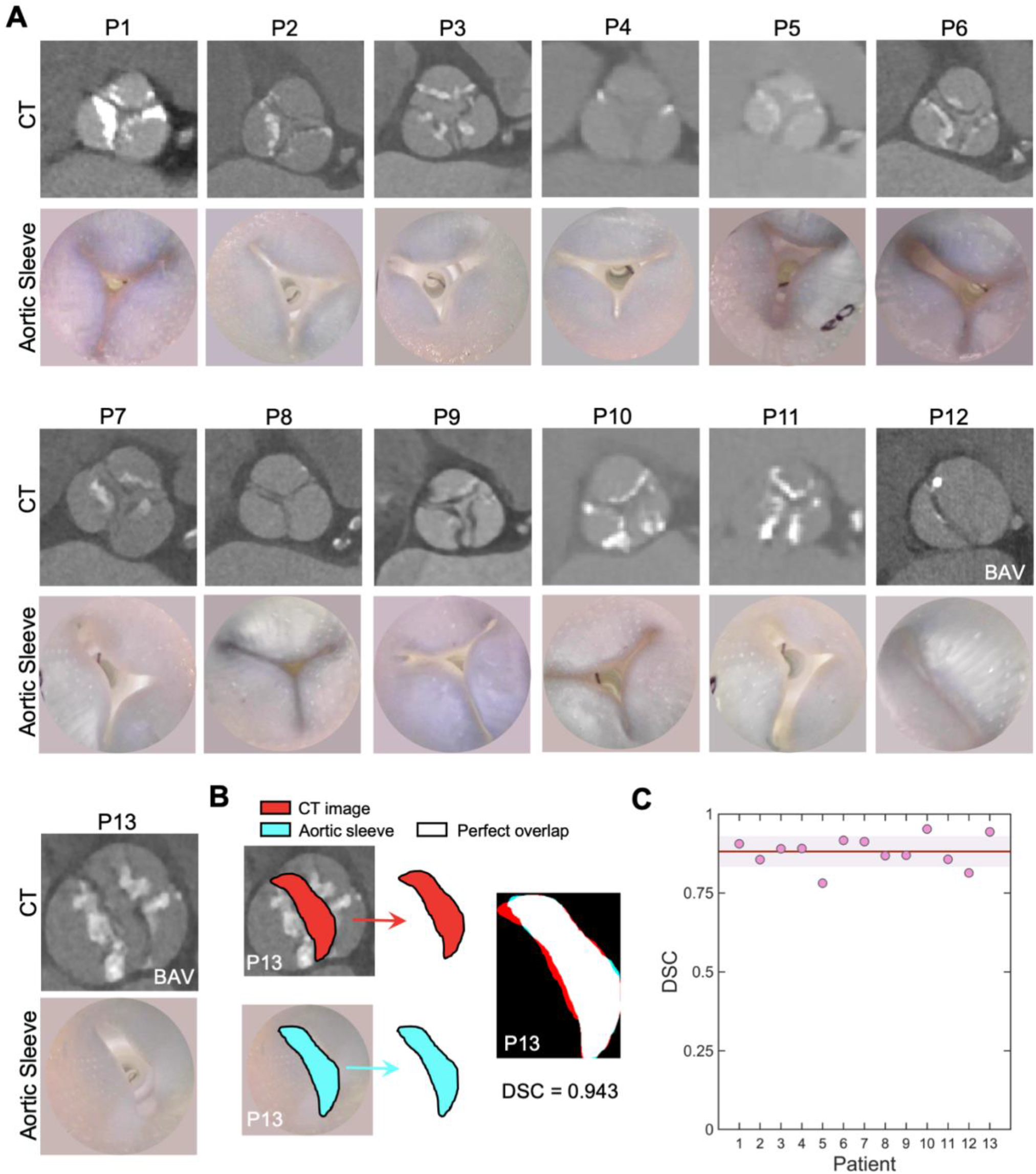
Morphologic mimicry of the aortic valve defect for both degenerative AS and BAV. (**A**) Top row: images of the aortic valve from CT data (patients 1-13). Bottom row: aortic lumen in the model during aortic sleeve actuation. All images taken at peak systole. (**B**) Representative CT and aortic sleeve luminal images (patient 13) for calculation of the DSC. (**C**) DSC for all patients. The red line in graph indicates the average DSC across the patient population and the shaded area shows ± 1 s.d. BAV: bicuspid aortic valve.

### Patient-specific aortic sleeves are tuned to recreate clinically relevant AS hemodynamics

By recreating patient-specific aortic valve anatomies, our soft robotic aortic sleeve was able to induce aortic valve hemodynamics as measured *via* TTE or TEE. We validated our model by comparing the clinical parameters measured in these patients for AS evaluation with those obtained by our system. We also compared our model with the MM3DP approach by Hosny *et al. (29)*, whereby CT images are used to generate 3D printed patient-specific aortic sinus and leaflet anatomies in tandem with rigid calcium-like patterns corresponding to the patients’ mineralized nodules. We improved the manufacturing approach by 3D printing the leaflets of the aortic valve and the rigid calcium-like nodules together with the patient-specific model of the LV and ascending aorta to perform functional hydrodynamic studies (fig. S5). Fig. 3A shows images of the soft robotic LV sleeve on the 3D printed anatomy, as well as the soft robotic aortic sleeve, and the MM3DP sinuses of the patient subgroup used for comparison.

**Fig. 3.**
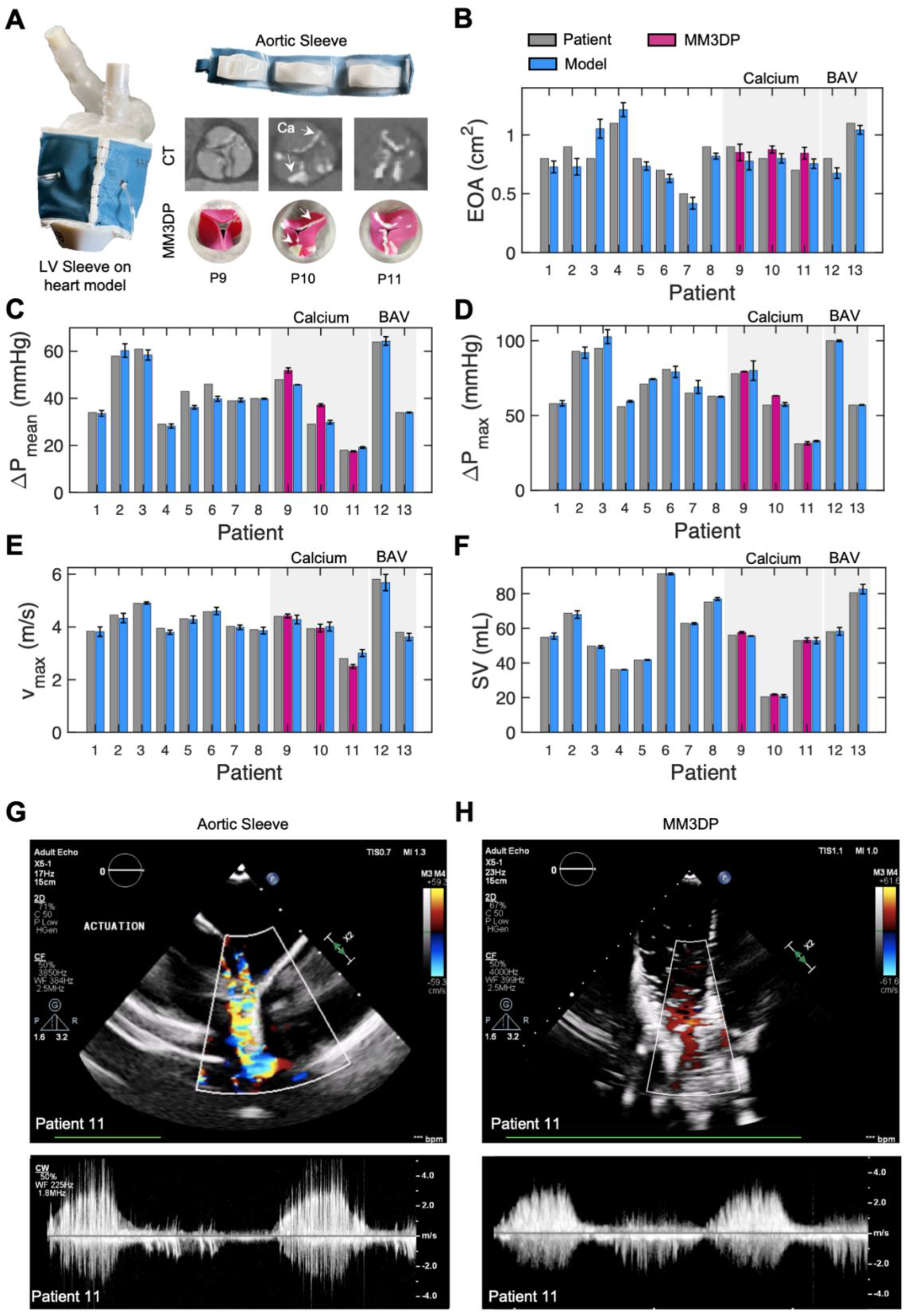
Soft robotic aortic sleeve recreates clinical metrics of AS. **(A**) Photo of the LV sleeve on the 3D printed heart model and of the soft robotic aortic sleeve. Photos of the patient-specific aortic valve leaflets and calcium (Ca) deposits manufactured using MM3DP technology for patients 9-11 and corresponding CT images. Measurements of (**B**) effective orifice area (EOA), (**C**) mean transaortic pressure gradient (ΔP_mean_), (**D**) maximum transaortic aortic pressure gradient (ΔP_max_), (**E)** peak aortic flow velocity (v_max_), and (**F**) stroke volume (SV) for each patient. Each graph shows a comparison between the hemodynamics measured in patients *via* TTE or TEE data, the values obtained in our model through actuation of the soft robotic sleeves, and the values obtained using MM3DP. Representative CFM and CW Doppler images for patient 11 obtained by (**G**) our model and (**H**) MM3DP, illustrating aortic velocity profiles characteristic of AS. Each bar indicates mean ± 1 s.d. (*n* = 3 consecutive heart cycles). BAV: bicuspid aortic valve.

Our model accurately recapitulated the effective orifice area (EOA; Fig. 3B) as well as the critical hemodynamic parameters of AS with high accuracy for each patient. For our analysis, we considered the mean (ΔP_mean_; Fig. 3C) and maximum transaortic pressure gradients (ΔP_max_; Fig. 3D), the peak aortic flow velocity (v_max_; Fig. 3E, fig. S6) and the stroke volume (SV; Fig. 3F). The variations from target values for each of these parameters were, on average, less than 5%, with - 4.3 ± 14.0 % for EOA, -2.2 ± 6.3 % for ΔP_mean_, 2.4 ± 3.5 % for ΔP_max_, -0.7 ± 3.1 % for v_max_, 0.3 ± 1.2 % for SV (*n* = 13).

Our soft robotic sleeve provides greater controllability through actuation modulation and higher fidelity morphologic and hemodynamic mimicry of AS than the MM3DP approach. With the MM3DP approach, variations from target values were 8.2 ± 13.0 %, 11.0 ± 15.9 %, 4.7 ± 5.3 %, - 3.2 ± 6.4 %, 3.2 ± 3.0 % for EOA, ΔP_mean_, ΔP_max_, v_max_, and SV respectively (*n* = 3). Finally, Fig. 3G-H shows color-flow mapping (CFM) Doppler images and corresponding aortic flow velocity tracings using continuous-wave (CW) Doppler obtained using the aortic sleeve (Fig. 3G) and MM3DP (Fig. 3H), illustrating the corresponding aortic velocity profiles.

### Dynamic tuning of soft robotic LV sleeve mimics changes in ventricular compliance secondary to AS

We customize the LV sleeve to the patient’s anatomy and actuated it to recreate anatomical filling, emptying, and wall motion during diastole (Fig 4A) and systole (Fig 4B). By tuning the actuation pressures of the LV sleeve during diastole, LV compliance and diastolic function can be modulated (Fig. 4C). This allows us to recreate elevations in the LV end-diastolic pressure (LVEDP; Fig. 4D) and in the end-diastolic pressure-volume relationship (EDPVR; Fig. 4E) associated with concentric remodeling secondary to AS.

**Fig. 4.**
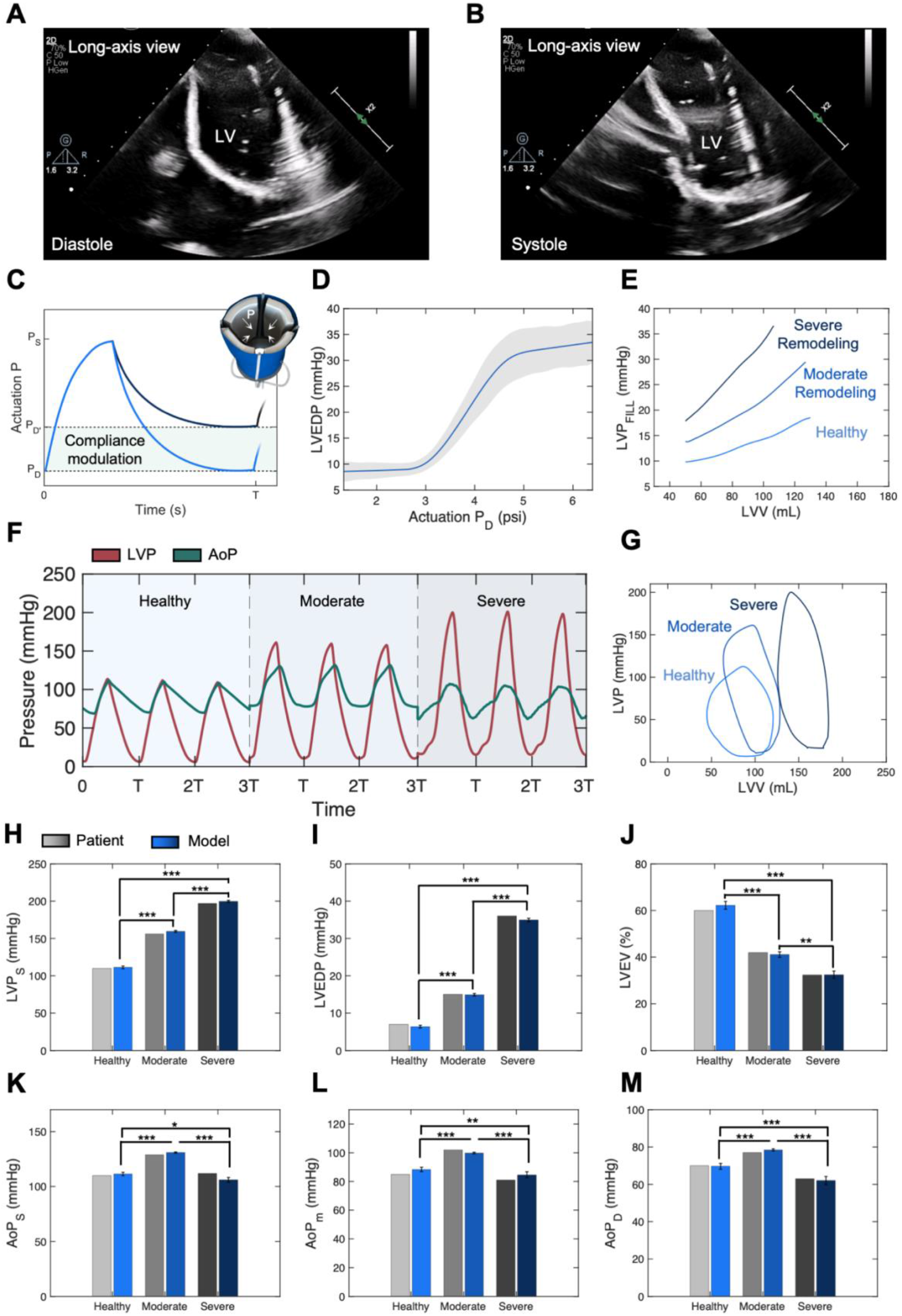
Patient-specific LV sleeve can be tuned to modulate ventricular elastance and simulate LV remodeling secondary to AS. Representative echocardiographic images of the soft 3D printed heart in long-axis view, during actuation by the soft robotic LV sleeve in (**A**) diastole and (**B**) systole. (**C**) Representation of the actuation pressure of the LV sleeve during an entire cardiac cycle, showing that end-diastolic pressure can be tuned to modulate ventricular compliance. P_S_: peak systolic actuation pressure. P_D_: end-diastolic actuation pressure. (**D**) Changes in left ventricular end-diastolic pressure (LVEDP) for various P_D_ values. (**E**) Representative end-diastolic pressure-volume relationship (EDPVR) curves for the simulated healthy LV, and moderate and severe concentric remodeling. LVP_FILL_: LV filling pressure; LVV: LV volume. (**F**) Representative superimposed LV (LVP) and aortic (AoP) pressure tracings for three consecutive heart cycles for the simulated healthy LV, and moderate and severe concentric remodeling. T: period of one heart cycle. (**G**) Sample LV pressure-volume (PV) loops for the simulated healthy LV, and moderate and severe concentric remodeling. Echocardiographic patient data and measured model values of the (**H**) left ventricular systolic pressure (LVP_S_), (**I**) LVEDP, (**J**) left ventricular ejection fraction (LVEF), (**K**) systolic aortic pressure (AoP_S_), (**L**) mean aortic pressure (AoP_m_), and (**M**) diastolic aortic pressure (AoP_D_) for the simulated healthy LV, and after moderate and severe concentric remodeling. Each bar indicates mean ± 1 s.d. (n = 3 consecutive heart cycles). Average values of healthy LV in (E-H) were taken from the literature *(7, 35)* and healthy waveforms were simulated using a functional bileaflet valve instead of the soft robotic aortic sleeve (E-H). Data from patients 11 and 12 were used as examples of moderate and severe remodeling, respectively. One-way ANOVA significance, *: *p* < 0.05; **: *p* < 0.01; ***: *p* < 0.001.

We simulated the hemodynamics of two patients that had LV catheterization data to demonstrate the ability of the soft robotic system to recreate cardiac hemodynamics of patients with different degrees of LV remodeling (patients 11 and 12). Specifically, patient 11 had mild AS (ΔP_max_ = 38 mmHg) and moderate changes in EDPVR (LVEDP = 15 mmHg), whereas patient 12 presented with severe AS (ΔP_mean_ = 64 mmHg) and a more severely elevated EDPVR (LVEDP = 36 mmHg) *(32, 34)*. For comparison, we recreated the hemodynamics of a healthy heart by implanting a functional surgical bileaflet valve in our system (fig. S5) and targeting typical healthy values as reported in the literature *(7, 35)*. Superimposed LV and aortic pressure tracings are displayed in Fig. 4F, with the corresponding PV loops shown in Fig. 4G. As expected, the transaortic pressure gradient, i.e., the difference between the LV and aortic pressure during systole, increases with the severity of AS (Fig. 4F), primarily driven by elevations in systolic LV pressures. Furthermore, LV filling pressures increase with severity of LV remodeling while SV decreases (Fig. 4G).

Measurements of the LV and aortic hemodynamics, i.e., systolic LV pressure (LVP_S_; Fig. 4H), LVEDP (Fig. 4I), LVEF (Fig. 4J), and systolic (AoP_S_; Fig. 4K), mean (AoP_m_; Fig. 4L), and diastolic (AoP_D_; Fig. 4M) aortic pressures, further corroborate the ability of our soft robotic aortic sleeve to create a high-fidelity model of each patients’ LV and aortic hemodynamics. For the two patients included in this analysis, we computed deviations from the corresponding target clinical values equal to 1.8 ± 0.6 % (LVP_S_), -1.7 ± 1.7 % (EDLVP), -0.7 ± 1.7 % (LVEF), -1.9 ± 4.8 % (AoP_S_), 1.2 ± 4.7 % (AoP_M_), 0.2 ± 2.4 % (AoP_D_) (*n* = 2). Comparing across the healthy and two diseased groups, there were statistically significant differences in the hemodynamic parameters evaluated (*p* < 0.05, Fig. 4 H-M). Overall, this approach showcases the ability of our platform to recreate LV hemodynamics and changes in LV compliance associated with remodeling processes secondary to AS with high fidelity.

### Soft robotic platform predicts the hemodynamic benefits of TAVR in patients with AS

We demonstrated potential use of our model in the evaluation of valvular prostheses in a subset of patients by comparing their hemodynamics prior to and following integration of a 26 mm Evolut R (Medtronic) self-expanding (SE) transcatheter valve in our soft robotic hydrodynamic system (fig. S5). Fig. 5A shows a schematic of the valve deployed through the distal end of the 3D printed anatomy and a detail of the valve outflow. For this study, we selected patients 4 and 11 based on the diameter of their aortic annulus (Table 1) and the manufacturer’s sizing guidelines *(36)*. We evaluated clinical parameters of AS for hemodynamic comparison pre- and post-implantation. In these patients, valve implantation resulted in an increase in EOA (Fig. 5B) by 95.0 ± 41.0 %, and a drop in ΔP_mean_ (Fig. 5C), ΔP_max_ (Fig. 5D), and v_max_ (Fig. 5E) by 83.5 ± 13.1 %, 73.9 ± 9.3 %, and 67.3 ± 12.4 %, respectively. Improvements in LV hemodynamics are visualized in Fig. 5F, which shows the LV PV loops of patient 11 pre- and post-valve implantation, highlighting a 34.7% drop in LVP_S_ and a 51.6% increase in SV.

**Fig. 5.**
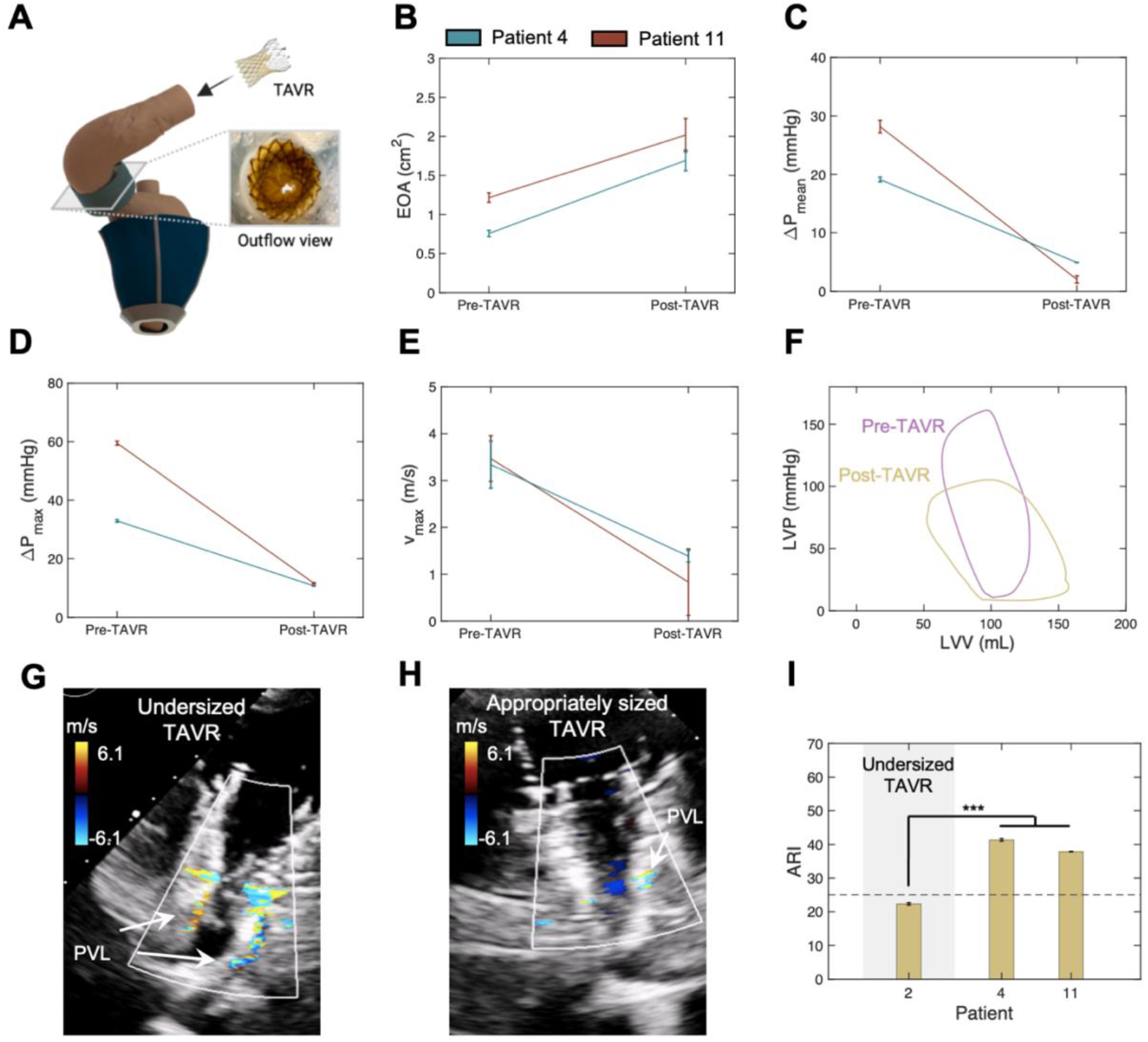
Usability of the model as a patient-specific testbed for the development and testing of devices for AS. (**A**) Illustration of the 3D printed heart, soft robotic sleeve, and of a transcatheter aortic valve replacement (TAVR) valve being inserted in the system, with a photo of the valve deployed in the aorta. Measurements of the (**B**) effective orifice area (EOA), (**C**) mean transaortic pressure gradient (ΔP_mean_), (**D**) maximum transaortic aortic pressure gradient (ΔP_max_), (**E)** peak aortic flow velocity (v_max_), and (**F**) LV pressure-volume (PV) loops for patient 11 before and after TAVR implantation. CFM Doppler images obtained during diastole highlighting the extent of paravalvular leakage (PVL) for the (**G**) undersized TAVR (patient 2) and (**H**) appropriately sized TAVR (patient 4) scenarios. (**I**) Aortic regurgitation index (ARI) of a patient with an undersized TAVR (patient 2) and two patients with an appropriately sized device (patients 4 and 11). Each plot indicates mean ± 1 s.d. (*n* = 3 consecutive heart cycles). *t*-test significance, *: *p* < 0.05; **: *p* < 0.01; ***: *p* < 0.001.

We used CFM Doppler to further evaluate valve sizing and regurgitation in the two different patient models. An appropriately sized valve (in patients 4 and 11) could be distinguished from a deliberately undersized valve (in patient 2) through assessment of paravalvular leakage (PVL) and the aortic regurgitation index (ARI). The undersized valve resulted in a greater degree of paravalvular leak on CFM Doppler than the appropriately sized valve (Fig. 5 G-H). Similarly, undersizing the valve resulted in an ARI lower than 25 compared to an average ARI of 39.6 ± 2.0 in the appropriately sized cases (Fig. 5I; *p*<0.001). This threshold of ARI 25 has been clinically associated with higher mortality *(37)*. Overall, this model was shown to enable evaluation of hemodynamic changes due to valve prostheses in various clinical scenarios.*(37, 38)(37)*

## DISCUSSION

In this work, we present the development of a patient-specific hydrodynamic model driven by tunable soft robotic tools, of relevance in AS and LV remodeling. We demonstrated the ability of this model to recreate patient-specific anatomies of degenerative stenotic aortic valves and of congenital BAV disease (Fig. 2A). Biomechanical mimicry of the AS lesion is paramount to accurately recreate local flow hemodynamics in a high-fidelity platform. In this work, calculation of the DSC between the model and CT images demonstrated that the patient-specific aortic sleeve can achieve enhanced mimicry (DSC = 0.88 ± 0.05) compared to commercial aortic banding techniques (DSC = 0.47) and non-specific aortic sleeve (DSC = 0.72) *(30)*. Furthermore, our platform recapitulates the hemodynamics of AS (Fig. 3 B-E) with greater accuracy compared to other systems, with a mean absolute deviation equal to 2.0% (n = 13), which is lower than that achieved by MMD3P techniques (5.2%, n = 3; Fig. 3 B-E).

The accuracy of molding- or MM3DP-based systems depends more heavily on the resolution of the patients’ CT images and on the quality of image segmentation than a soft robotics-driven platform. In the aforementioned systems, any mismatch between the patients’ anatomies and the aortic root replicas can only be improved by manual editing of the digital valvular geometries. This time-consuming iterative process compromises the utility of these models in the clinical setting, where TAVR procedures are preferably performed within a day from CT imaging. Conversely, in a soft robotic model, actuation can be tuned in real-time to obtain high-fidelity mimicry of the patients’ hemodynamics, by modulating the aortic root diameter (Fig. 3G). The controllability of molding or MM3DP models is further limited by differences between the mechanical properties of the valve material and those of the native leaflet tissue. Although the investigation of valve kinetics was beyond the scope of this work, the dynamics of the aortic sleeve in this model could be controlled to mimic the motion of stenotic valve leaflets, as previously demonstrated *(30)*. Furthermore, unlike other models, our platform was able to recreate the anatomies of congenital valvular defects, such as BAV (patients 12-13 in Fig. 2A and Fig. 3B-F), which is a primary driver of AS in the younger population *(3)*. The MM3DP approach developed by *Hosny* et al. *(29)* relies on an algorithm that computes an idealized geometry of a tricuspid aortic valve from CT image landmarks, and is yet to be broadened to recreate BAV anatomies or other congenital aortic valve defects.

Integration of a controllable soft robotic LV sleeve (Fig. 3A, Fig. 4A-B) is a critical step towards the development of clinically relevant hydrodynamic models of AS and other cardiovascular conditions. Traditional hydrodynamic models leverage displacement-control pumps, which eject a prescribed amount of volume into the circulation. First, this is not representative of cardiac physiology; second, it makes it challenging to recreate the hemodynamics of conditions where the afterload is altered. In the context of AS, these systems would not be able to capture drops in SV associated with a higher afterload (Fig. 3F), leading to overestimated values of flow velocities predicted by the continuity equation (Fig. 3E) *(39, 40)*. Conversely, by mimicking the biomechanics of the native heart, we overcome these limitations and recreate both pressure and flow in a more physiological manner (Fig. 4 F-G) *(41–44)*.

The personalized LV sleeve design enables us to modulate LV compliance to simulate the hemodynamic effects of cardiac remodeling secondary to AS in a patient-specific manner (Fig. 4C). Particularly, we were able to simulate alterations in LV filling pressures in patients with moderate and severe remodeling due to pressure overload (Fig. 4D-E). This model of modulation of ventricular compliance can be used to represent different states of disease progression, which has not been shown previously. As thickening and subsequent decrease of LV compliance are estimated to occur in more than two-thirds of patients with AS *(8)*, it is paramount that preclinical models of AS can correctly recapitulate changes in LV diastolic biomechanics and hemodynamics associated with pressure overload.

It is worth noting that there are three main limitations associated with our study. First, the position of the aortic sleeve on the ascending aorta may affect the compliance of the 3D printed model. Particularly, the aortic sleeve causes a drop in distension of the aortic segment corresponding to the position of the sleeve. In addition, any aortic segments that are proximal to the aortic sleeve will experience LV (rather than aortic) pressures, which are elevated in AS. Altogether, these factors may lead to local differences in aortic distension compared to physiology. Second, although our model was shown to capture LV pressures and flows (Fig. 4 F-G) with high accuracy compared to other hydrodynamic models *(45–47)*, the isovolumic regions of LV PV loops display a non-zero net flow towards or outside of the LV. This is a result of the position of the aortic flow probe, which could only be placed distal to the aortic valve plane due to the irregular geometry of the 3D printed aortic anatomies. Finally, the dataset used in this study did not provide indications of the patients’ systolic-diastolic ratio, which limited our ability to modulate the exact dynamics of ventricular contraction and may have impacted hemodynamic measurements.

Despite these limitations, this work has the potential to shift the paradigm for device development in the industrial setting and for procedural planning in the clinic, as suggested by our preliminary TAVR use investigation (Fig. 5). This research could enable medical device companies to test and optimize their devices reliably across a spectrum of clinical cases, broadening the usability of devices to those patients for whom current TAVR designs are not suitable, beneficial, or safe to use. In the clinic, it would provide physicians with a platform for device selection, and procedural planning and outcome prediction. Furthermore, it may provide clinicians with a tool to improve techniques for TAVR delivery to minimize risk of coronary obstruction or valve migration and optimize device selection for patients with complex anatomies or sizes that fall between recommended use ranges for a given device. Finally, it may help identify subgroups for which TAVR could be the beneficial and performed safely within patient populations - such as BAV patients - that are currently ineligible for TAVR and have been traditionally excluded from major trials comparing surgical versus transcatheter interventions *(48, 49)*.

In future work, we aim to automate the sleeve design process and improve current 3D printing techniques to further reduce the manufacturing time of the patients’ replicas - currently estimated to be approximately 20 hours - to maximize clinical utility. Further, we hope to obtain access to a broader spectrum of transcatheter valve prostheses and a larger clinical database, including post-TAVR hemodynamic measurements, for validation of our TAVR prediction study. In addition, we aim to conduct prospective clinical trials to support physicians in their TAVR device selection process and prospectively validate use of our model.

Eventually, use of this soft robotics-driven model can be broadened to simulate the hemodynamics of other valvular heart diseases and conditions that affect LV function, including restrictive cardiomyopathies and heart failure – both with reduced (HFrEF) and preserved ejection fraction (HFpEF). We are hopeful that this model can pave the way towards high-fidelity patient-specific tunable models with a translational potential poised to revolutionize clinical care of the millions of people worldwide affected by AS and other cardiovascular conditions.

## MATERIALS AND METHODS

### Study design

Anonymized CT and echocardiographic clinical data were obtained retrospectively *via* IRB approval at the Massachusetts General Hospital. Using echocardiographic measurements of the left ventricular (LV) diameter during systole and diastole, we screened for chest CT images acquired during diastole. This allowed us to design the geometry of our patient-specific soft robotic LV sleeve in its pre-actuation state. Conversely, images of the aortic valve were taken during peak systole from the patients’ aortic valve cine images. This enabled us to develop patient-specific soft robotic aortic sleeve that could recreate the morphology of the stenotic leaflets during systole. Each patient’s 3D printed anatomical model was integrated with the LV and aortic sleeves into a hydrodynamic flow loop, and added pressure and flow sensors, and an endoscopic camera for hemodynamic evaluation. Hemodynamic parameters relevant in AS were measured using pressure-volume catheters, flow probes, an endoscopic camera, and CW and CFM Doppler, as described below. Results were compared with the patients’ clinical data, as well as established methods based on MM3DP. Finally, hemodynamic changes due to implantation of a 26mm self-expanding (SE) Evolut R transcatheter aortic valve prosthesis (Medtronic) were evaluated.

### Patient CT data segmentation and 3D printing of cardiac and aortic vascular anatomies

CT and aortic valve cine images (slice thickness t = 1.0-1.5 mm, x-ray depth D = 10 mm) were segmented on Mimics Research software (v.21.0.0.406, Materialise, NV) by thresholding, multiple-slice editing, and auto-interpolation. The geometrical axes of the cine images were reoriented to ensure visualization of the valve leaflets orthogonal to the direction of flow.

For each patient, the 3D anatomy of the LV and aorta (ascending through descending segment) was exported from the CT images as a shell stereolithography (.stl) file with wall thickness equal to 1.3 mm. A thickness value lower than that of the human aorta was chosen to compensate for any mismatch in mechanical properties between the 3D printing photopolymer resin (Elastic 50A; Formlabs, Somerville, MA) and those of the native aorta (fig. S1). The mechanics of the LV are defined by actuation pressures of the LV sleeve; therefore, a thickness value of 1.3mm for the 3D printed LV wall was chosen for ease of manufacturing. Each .stl file was then imported to Preform software (v 3.21 or above, Formlabs) and the architecture of the support material was manually adjusted to avoid any overhang and to minimize the presence of internal support material. Each anatomy was then printed on a Form 3B Stereolithography (SLA) 3D-printer (Formlabs Inc., Somerville, MA) with a layer thickness of 0.1 mm.

### LV and aortic sleeve design and manufacturing

Each anatomical (.stl) LV model was used for the design of a patient-specific soft robotic LV sleeve on SOLIDWORKS (Dassault Systèmes, 2019). The outer surface of the LV was offset by 10 mm to generate guide tracings for the sleeve geometry and divided the tracings into four circumferential quadrants (i.e., each approximately 90 degrees apart) and. These quadrants were flattened to a plane to create the contours of the molds for manufacturing. The flat tracings were then extruded by the same offset (10 mm) and 3D printed each mold using a rigid material (i.e., Veroblue, Stratasys) on a Fused Deposition Modeling (FDM) 3000 Objet 3D-printer (Stratasys). Similarly, the contours of the aortic valve leaflets were exported from CT images and converted into flat geometries, which were then extruded and 3D printed for manufacturing of the aortic sleeve.

Analogously to the manufacturing technique previously described by our group *(30, 50)*, for each of the four LV molds and three aortic molds (or two for bicuspid valve anatomies) per patient, two sheets of Thermoplastic Polyurethane (TPU, HTM 8001-M 80A shore polyether film, 0.012” thick, American Polyfilm, Inc) were vacuum-formed (Yescom Dental Vacuum Former, Generic) into the shape of the molds. Each pair of TPU was then heat-sealed at 320F for 8 seconds on a heat press transfer machine (Heat Transfer Machine QXAi, Powerpress) using negative acrylic molds to create enclosed and inflatable geometries. For each sleeve, these inflatable pockets were then heat-sealed using a similar process as that described above to a 200-Denier TPU-coated fabric (Oxford fabric, Seattle fabrics Inc.), which was designed to fit around their respective LV or aortic anatomy. Further, holes were opened through the fabric on one side of each of the pockets to connect soft tubes (latex rubber 1/16” ID 1/8” OD tubing, McMaster-Carr) as actuation lines through PVC connectors (polycarbonate plastic double-barbed tube fitting for 1/16” tube ID, McMaster-Carr).

A 3D printed (3000 Objet, Stratasys) rigid skeleton with four arms (one for each pair of adjacent pockets of the LV sleeve) was designed to secure each LV sleeve to the corresponding 3D printed geometry. A belt-like securing mechanism with Velcro-adhesive was integrated in the design of the fabric of each aortic sleeve to guarantee secure attachment around the 3D printed aorta (fig. S2).

### 3D printing of calcified valves

In this work, the multi-material 3D printing approach developed by Hosny *et al (29)* was optimized for hydrodynamic testing and used for comparison with our model. We used the algorithm developed in their work to generate the aortic valve leaflet geometries from landmarks of abdominal CT images with superimposed calcium-like nodules. However, instead of limiting the vascular anatomy to the aortic sinus, we integrated the valve leaflets and nodules into the entire LV and aortic (through the aortic arch segment) anatomies. First, this allowed us to conduct functional tests of their model of AS; second, it enabled us to integrate their approach with our strategy of LV actuation, allowing for a fairer comparison between the two models of AS. The LV, aorta, aortic valve leaflets, and calcium nodules were printed simultaneously using an Object 500 Connex 3D-printer (Stratasys) using the same printing techniques as what are described in the original paper *(29)*. To do this, we created a small (2-5 mm in diameter) hole in proximity to the LV apex to remove any support material laid down during the manufacturing process. The hole was then sealed using the same resin material and UV light was applied manually.

### Patient-specific hydrodynamic studies

For each patient, a closed loop was set up for hydrodynamic studies (viscosity of medium, μ =1,0 cP) *(21)*. The loop was assembled by connecting the 3D printed anatomy to a series of soft PVC plastic tubing (3/8”-5/8” ID, 5/8-1” OD, McMasterr-Carr), two variable-resistance ball valves to mimic arterial and venous resistance, and two custom-made acrylic compliance chambers to recreate peripheral compliance. A unidirectional mechanical valve (Regent bileaflet mechanical prosthesis, 19AGN-751 standard cuff, Abbott) was connected in proximity to the venous system.

Two clamp-on perivascular flow probes (PS series, Transonic) were used to measure flow immediately distal to the descending aorta (LV outflow) and distal to the surgical valve in the LV (LV inflow). The flow probes were connected to a two-channel flowmeter console (400-series, Transonic), which was in turn connected to an 8-channel Powerlab system (ADInstruments) for data acquisition and recording. Two straight-tip 5F pressure-volume (PV) catheters were inserted through two adjustable catheter connectors to measure pressures at the LV and at the ascending aorta, distal to the aortic sleeve. The catheters were connected to a Transonic ADV500 PV System and to the Powerlab (ADInstruments). Given the mismatch in electrical impedance between the 3D printing material and that of the native cardiac tissue, the PV catheters could not be used to measure volumes inside the LV. An endoscopic camera (1080P HD, 30 fps, NIDAGE) was inserted in the system to visualize the cross-sectional profiles of the aorta during actuation for subsequent calculations of the valve EOA.

The system was actuated pneumatically through the soft robotic LV sleeve, which was connected to a control box and associated GUI, where input pressure tracings could be defined (Fig. 4C). The aortic sleeve was actuated hydraulically using a syringe pump (70-3007 PHD ULTRA™ Syringe Pump Infuse/Withdraw, Harvard Apparatus). The actuation pressures and volumes of the soft robotic sleeves were modulated to achieve the values of SV and ΔP_max_ for each individual patient, as well as LV and aortic pressure values when known. Systolic and diastolic actuation pressures of the LV sleeves ranged between 8-13 psi and 0-6 psi for systole and diastole, respectively; whereas actuation volumes equal to 20 – 40 mL were used for actuation of the aortic sleeves.

### Echocardiographic evaluation

The Epiq CVx cardiovascular ultrasound system (Philips) was used in tandem with the X5-1 transducer (Philips) for echocardiographic evaluation of each patient-specific model to assess the degree of AS and, in some instances, LV function and PVL. As the transaortic pressure gradients and the EOA could be more accurately measured using the PV catheters and the endoluminal camera respectively, echocardiography was primarily used to compute the peak flow velocity (v_max_) through the aortic valve (fig. S6). The probe was positioned directly on the 3D printed geometry, leveraging the anatomical curvature between the ascending aorta and proximal arch to align the ultrasonic beam with the direction of flow for CW Doppler imaging. Tracings of the aortic flow velocity were obtained, and the peak value of each tracing (v_max_) was calculated. In a subset of patient models, CFM Doppler images were obtained for visualization of the flow through the soft robotic aortic sleeve and stenotic MM3DP valves for comparison. In addition, 2D movies of the LV in long-axis view, of the MM3DP valve, and of the SE valve prosthesis (Evolut R, Medtronic) were recorded to visualize actuation of the soft robotic LV sleeve (Fig. 4A-B) and mobility of the MM3DP and TAVR leaflets. Finally, CFM Doppler images were taken to provide a qualitative comparison of the degree of PVL between the patient models with an appropriately sized and an undersized valve (Fig. 5 G-H).

### Evaluation of post-TAVR hemodynamics

A 26-mm Evolut R (Medtronic) self-expanding (SE) transcatheter aortic valve prosthesis was implanted in a total of three patient models (patients 2, 4, and 11). The valve was delivered manually from a distal opening in the anatomy to the point of constriction or slightly supra-annularly. We used a valve that was undersized for patient 2 and appropriately sized for the other patients, based on the patients’ annular diameters and the manufacturer’s sizing specifications. This enabled us to characterize hemodynamic benefits in these patients. Additionally, through calculations of the aortic regurgitation index (ARI) and CFM Doppler imaging, we could visualize and compare the degree of regurgitation in these patient groups. The ARI was calculated as per Equation (1):

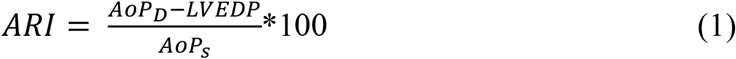

where AoP_D_ and AoP_S_ are the diastolic and systolic aortic pressures, respectively, and the LVEDP is the end-diastolic LV pressure. All parameters in this equation were measured from PV catheters.

### Data analysis

Data were primarily visualized and acquired by LabChart (Pro v8.1.16, ADInstruments). All the input signals were filtered using the default 50Hz band-stop filter. Signals include the LV and aortic pressures for calculation of the transaortic pressure gradient, the flow rates out of and into the LV for calculation of the stroke volume (SV) and ejection fraction (LVEF). From these data, the peak, mean, and end-diastolic LV and aortic pressures were extracted, and the LVP and aortic pressures were plotted over time and used for calculation of the PV loops, when clinical PV reports were available. Analogously, the actuation pressure of the LV sleeve was displayed and recorded on LabChart. Data analysis and visualization was performed through an automated algorithm developed on MATLAB R2020a (MathWorks). Values for peak flow velocity measured using CW Doppler were included in the analysis. Average values and standard deviations were calculated for three consecutive heart cycles after the soft robotic sleeve actuation degrees were successfully tuned to recreate the patients’ cardiac and aortic hemodynamics. Where applicable, one-way analysis of variance (ANOVA) and t-tests were performed (MATLAB R2020a, MathWorks) to determine significance between groups. P-values were adjusted for multiple comparison as appropriate.

Images of the aortic cross-sections were processed using the Image Processing and Computer Vision MATLAB toolbox (Mathworks) for calculation of the EOA. These images and those obtained from the patients’ CT were binarized and each pair of images (one pair per patient) was cross-registered. The *rigid* distortion option i.e., enabling only translation and rotation of the moving image, was utilized for image registration, and the Sorensen-Dice similarity coefficient (DSC) was calculated for each patient model.

## Data Availability

All data produced in the present study are available upon reasonable request to the authors

## List of Supplementary Materials

Table S1

Fig. S1to S6

Movie S1 to S3

## Author contributions

Conceptualization: LR, CO, CTN, ETR

Methodology: LR, CO, DG, JB, SXW, BB, JW, CTN, ETR

Investigation: LR, CO, DG, JB, SXW, CTN, ETR

Visualization: LR

Funding acquisition: ETR

Project administration: CTN, ETR Supervision: CTN, ETR

Writing – original draft: LR

Writing – review & editing: LR, CO, DG, JB, SXW, CTN, ETR

## Data and materials availability

All data and analysis code are available upon reasonable request.

## References

1. G. A. Fishbein, M. C. Fishbein, Pathology of the Aortic Valve: Aortic Valve Stenosis/Aortic RegurgitationCurr. Cardiol. Rep. 21 (2019), doi:10.1007/s11886-019-1162-4.

2. J. Ternacle, L. Krapf, D. Mothy, J. Magne, A. Nguyen, A. Galat, R. Gallet, E. Teiger, N. Côté, M. A. Clavel, F. Tournoux, P. Pibarot, T. Damy, Aortic Stenosis and Cardiac Amyloidosis: JACC Review Topic of the WeekJ. Am. Coll. Cardiol. 74, 2638–2651 (2019).

3. W. C. Roberts, J. M. Ko, Frequency by decades of unicuspid, bicuspid, and tricuspid aortic valves in adults having isolated aortic valve replacement for aortic stenosis, with or without associated aortic regurgitation, Circulation 111, 920–925 (2005).

4. R. Rajani, J. Hancock, J. B. Chambers, The art of assessing aortic stenosisHeart 98 (2012), doi:10.1136/heartjnl-2012-302392.

5. B. A. Carabello, W. J. Paulus, Aortic stenosisLancet 373, 956–966 (2009).

6. M. Gotzmann, S. Hauptmann, M. Hogeweg, D. S. Choudhury, F. Schiedat, J. W. Dietrich, T. H. Westhoff, M. Bergbauer, A. Mügge, Hemodynamics of paradoxical severe aortic stenosis: insight from a pressure–volume loop analysis, Clin. Res. Cardiol. 108, 931–939 (2019).

7. L. Rosalia, C. Ozturk, D. Van Story, M. A. Horvath, E. T. Roche, Object-Oriented Lumped-Parameter Modeling of the Cardiovascular System for Physiological and Pathophysiological Conditions, Adv. Theory Simulations 4, 2000216 (2021).

8. M. R. Dweck, S. Joshi, T. Murigu, A. Gulati, F. Alpendurada, A. Jabbour, A. Maceira, I. Roussin, D. B. Northridge, P. J. Kilner, S. A. Cook, N. A. Boon, J. Pepper, R. H. Mohiaddin, D. E. Newby, D. J. Pennell, S. K. Prasad, Left ventricular remodeling and hypertrophy in patients with aortic stenosis: insights from cardiovascular magnetic resonance, (2012), doi:10.1186/1532-429X-14-50.

9. L. Rosalia, C. Ozturk, S. Shoar, Y. Fan, G. Malone, F. H. Cheema, C. Conway, R. A. Byrne, G. P. Duffy, A. Malone, E. T. Roche, A. Hameed, Device-Based Solutions to Improve Cardiac Physiology and Hemodynamics in Heart Failure With Preserved Ejection Fraction, JACC Basic to Transl. Sci. (2021), doi:10.1016/J.JACBTS.2021.06.002.

10. M. A. Pfeffer, in Atlas of Heart Failure, W. S. Colucci, Ed. (Current Medicine Group, London, 2002), pp. 87–101.

11. F. Rader, E. Sachdev, R. Arsanjani, R. J. Siegel, Left Ventricular Hypertrophy in Valvular Aortic Stenosis: Mechanisms and Clinical Implications, Am. J. Med. 128, 344–352 (2015).

12. G. Barletta, M. R. Del Bene, F. Venditti, G. Pilato, P. Stefàno, Surgical aortic valve replacement and left ventricular remodeling: Survival and sex-related differences, Echocardiography 38, 1095–1103 (2021).

13. A. I. Duncan, B. S. Lowe, M. J. Garcia, M. Xu, A. M. Gillinov, T. Mihaljevic, C. G. Koch, Influence of concentric left ventricular remodeling on early mortality after aortic valve replacement, Ann. Thorac. Surg. 85, 2030–2039 (2008).

14. J. Joseph, S. Y. Naqvi, J. Giri, S. Goldberg, Aortic Stenosis: Pathophysiology, Diagnosis, and Therapy, Am. J. Med. 130, 253–263 (2017).

15. G. H. Bevan, D. A. Zidar, R. A. Josephson, S. G. Al-Kindi, Mortality Due to Aortic Stenosis in the United States, 2008-2017, JAMA 321, 2236–2238 (2019).

16. G. S. Gheorghe, A. S. Hodorogea, A. C. Dan Gheorghe, I. T. Nanea, A. Ciobanu, Medical management of symptomatic severe aortic stenosis in patients non-eligible for transcatheter aortic valve implantation, J. Geriatr. Cardiol. 17, 704 (2020).

17. A. Mazine, M. Ouzounian, Aortic valve replacement in young and middle-aged adults: looking beyond the tree that hides the forest, Ann. Transl. Med. 5 (2017), doi:10.21037/ATM.2017.02.06.

18. Research and Markets, Transcatheter Heart Valve Replacement Market, Global Forecast 2022-2027, Industry Trends, Impact of COVID-19, Opportunity Company Analysis (2022).

19. R. Puri, C. Chamandi, T. Rodriguez-Gabella, J. Rodés-Cabau, Future of transcatheter aortic valve implantation — evolving clinical indications, Nat. Rev. Cardiol. 2017 151 15, 57–65 (2017).

20. A. Rosenzweig, The Growing Importance of Basic Models of Cardiovascular Disease., Circ. Res. 130, 1743–1746 (2022).

21. International Organization for Standardization (ISO), ISO - ISO 5840-1:2021 - Cardiovascular implants — Cardiac valve prostheses — Part 1: General requirements (2021; https://www.iso.org/standard/77033.html).

22. P. Haaf, M. Steiner, T. Attmann, G. Pfister, J. Cremer, G. Lutter, A novel pulse duplicator system: evaluation of different valve prostheses, Thorac. Cardiovasc. Surg. 57, 10–17 (2009).

23. R. Medero, S. García-Rodríguez, C. J. François, A. Roldán-Alzate, Patient-specific in vitro models for hemodynamic analysis of congenital heart disease - Additive manufacturing approach, J. Biomech. 54, 111–116 (2017).

24. D. Chen, S. Liang, Z. Li, Y. Mei, H. Dong, Y. Ma, J. Zhao, S. Xu, J. Zheng, J. Xiong, A Mock Circulation Loop for In Vitro Hemodynamic Evaluation of Aorta: Application in Aortic Dissection, J. Endovasc. Ther. 29, 132–142 (2022).

25. M. Bonfanti, G. Franzetti, S. Homer-Vanniasinkam, V. Díaz-Zuccarini, S. Balabani, A Combined In Vivo, In Vitro, In Silico Approach for Patient-Specific Haemodynamic Studies of Aortic Dissection, Ann. Biomed. Eng. 48, 2950–2964 (2020).

26. A. Roldán-Alzate, S. García-Rodríguez, P. V. Anagnostopoulos, S. Srinivasan, O. Wieben, C. J. François, Hemodynamic study of TCPC using in vivo and in vitro 4D Flow MRI and numerical simulation, J. Biomech. 48, 1325–1330 (2015).

27. B. J. Kovarovic, O. M. Rotman, P. Parikh, M. J. Slepian, D. Bluestein, Patient-specific in vitro testing for evaluating TAVR clinical performance-A complementary approach to current ISO standard testing, Artif. Organs 45, E41–E52 (2021).

28. G. Haghiashtiani, K. Qiu, J. D. Z. Sanchez, Z. J. Fuenning, P. Nair, S. E. Ahlberg, P. A. Iaizzo, M. C. McAlpine, 3D printed patient-specific aortic root models with internal sensors for minimally invasive applications, Sci. Adv. 6, 4641–4669 (2020).

29. A. Hosny, J. D. Dilley, T. Kelil, M. Mathur, M. N. Dean, J. C. Weaver, B. Ripley, Preprocedural fit-testing of TAVR valves using parametric modeling and 3D printing, J. Cardiovasc. Comput. Tomogr. 13, 21–30 (2019).

30. L. Rosalia, C. Ozturk, S. A. Chen, R. A. Eder, J. A. Kim, M. Panagia, J. Guerrero, E. T. Roche, C. Nguyen, Recapitulating aortic valve disease hemodynamics with a highly tunable bioinspired soft robotic aortic sleeve, Res. Sq. (2021), doi:10.21203/rs.3.rs-713582/v1 (pre-print).

31. P. A. Heidenreich, B. Bozkurt, D. Aguilar, L. A. Allen, J. J. Byun, M. M. Colvin, A. Deswal, M. H. Drazner, S. M. Dunlay, L. R. Evers, J. C. Fang, S. E. Fedson, G. C. Fonarow, S. S. Hayek, A. F. Hernandez, P. Khazanie, M. M. Kittleson, C. S. Lee, M. S. Link, C. A. Milano, L. C. Nnacheta, A. T. Sandhu, L. W. Stevenson, O. Vardeny, A. R. Vest, C. W. Yancy, 2022 AHA/ACC/HFSA Guideline for the Management of Heart Failure: A Report of the American College of Cardiology/American Heart Association Joint Committee on Clinical Practice Guidelines, Circulation 145, E895–E1032 (2022).

32. C. M. Otto, R. A. Nishimura, R. O. Bonow, B. A. Carabello, J. P. Erwin, F. Gentile, H. Jneid, E. V Krieger, M. Mack, C. McLeod, P. T. O’Gara, V. H. Rigolin, T. M. Sundt, A. Thompson, C. Toly, 2020 ACC/AHA Guideline for the Management of Patients With Valvular Heart Disease: A Report of the American College of Cardiology/American Heart Association Joint Committee on Clinical Practice Guidelines, Circulation 143, e72–e227 (2021).

33. K. H. Zou, S. K. Warfield, A. Bharatha, C. M. C. Tempany, M. R. Kaus, S. J. Haker, W. M. Wells, F. A. Jolesz, R. Kikinis, Statistical Validation of Image Segmentation Quality Based on a Spatial Overlap Index: Scientific Reports, Acad. Radiol. 11, 178 (2004).

34. W. J. Paulus, C. Tschöpe, J. E. Sanderson, C. Rusconi, F. A. Flachskampf, F. E. Rademakers, P. Marino, O. A. Smiseth, G. De Keulenaer, A. F. Leite-Moreira, A. Borbély, I. Édes, M. L. Handoko, S. Heymans, N. Pezzali, B. Pieske, K. Dickstein, A. G. Fraser, D. L. Brutsaert, in European Heart Journal, (2007), vol. 28, pp. 2539–2550.

35. L. S. Lilly, Pathophysiology of heart disease: A collaborative project of medical students and faculty: Fifth edition L. S. Lilly, Ed. (Wolters Kluwer/Lippincott Williams & Wilkins, Baltimore, MD, ed. 6, 2013).

36. J. M. Sinning, N. Werner, G. Nickenig, E. Grube, Medtronic CoreValve Evolut valve, EuroIntervention 8, Q94–Q96 (2012).

37. J. M. Sinning, C. Hammerstingl, M. Vasa-Nicotera, V. Adenauer, S. J. Lema Cachiguango, C. Scheer, S. Hausen, A. Sedaghat, A. Ghanem, C. Mller, E. Grube, G. Nickenig, N. Werner, Aortic Regurgitation Index Defines Severity of Peri-Prosthetic Regurgitation and Predicts Outcome in Patients After Transcatheter Aortic Valve Implantation, J. Am. Coll. Cardiol. 59, 1134–1141 (2012).

38. W. A. Zoghbi, F. M. Asch, C. Bruce, L. D. Gillam, P. A. Grayburn, R. T. Hahn, I. Inglessis, M. Islam, S. Lerakis, S. H. Little, R. J. Siegel, N. Skubas, T. C. Slesnick, W. J. Stewart, P. Thavendiranathan, N. J. Weissman, S. Yasukochi, K. G. Zimmerman, Guidelines for the Evaluation of Valvular Regurgitation After Percutaneous Valve Repair or Replacement: A Report from the American Society of Echocardiography Developed in Collaboration with the Society for Cardiovascular Angiography and Interventions, Japanese Society of Echocardiography, and Society for Cardiovascular Magnetic Resonance, J. Am. Soc. Echocardiogr. 32, 431–475 (2019).

39. R. A. Migliore, M. E. Adaniya, M. Barranco, G. Miramont, S. Gonzalez, H. Tamagusuku, R. Migliore, M. E. Adaniya, M. Barranco, G. Miramont, S. Gonzalez, H. Tamagusuku, Estimation of Contraction Coefficient of Gorlin Equation for Assessment of Aortic Valve Area in Aortic Stenosis, World J. Cardiovasc. Dis. 7, 119–130 (2017).

40. R. Taylor, Evolution of the continuity equation in the Doppler echocardiographic assessment of the severity of valvular aortic stenosis, J. Am. Soc. Echocardiogr. 3, 326–330 (1990).

41. E. T. Roche, M. A. Horvath, I. Wamala, A. Alazmani, S. E. Song, W. Whyte, Z. Machaidze, C. J. Payne, J. C. Weaver, G. Fishbein, J. Kuebler, N. V. Vasilyev, D. J. Mooney, F. A. Pigula, C. J. Walsh, Soft robotic sleeve supports heart function, Sci. Transl. Med. 9 (2017), doi:10.1126/scitranslmed.aaf3925.

42. E. T. Roche, R. Wohlfarth, J. T. B. Overvelde, N. V. Vasilyev, F. A. Pigula, D. J. Mooney, K. Bertoldi, C. J. Walsh, A Bioinspired Soft Actuated Material, Adv. Mater. 26, 1200–1206 (2014).

43. L. Rosalia, E. Roche, M. Y. Saeed, in Advances in cardiovascular technology, J. H. Karimov, K. Fukamachi, M. Gillinov, Eds. (Elsevier, 2022).

44. J. Bonnemain, P. J. del Nido, E. T. Roche, Direct Cardiac Compression Devices to Augment Heart Biomechanics and Function, https://doi.org/10.1146/annurev-bioeng-110220-025309 24, p137–156 (2022).

45. K. Fukamachi, D. J. Horvath, J. H. Karimov, Y. Kado, T. Miyamoto, B. D. Kuban, R. C. Starling, Left atrial assist device to treat patients with heart failure with preserved ejection fraction: Initial in vitro study, J. Thorac. Cardiovasc. Surg. (2020), doi:10.1016/j.jtcvs.2019.12.110.

46. C. Miyagi, B. D. Kuban, C. R. Flick, A. R. Polakowski, T. Miyamoto, J. H. Karimov, R. C. Starling, K. Fukamachi, Left atrial assist device for heart failure with preserved ejection fraction: initial results with torque control mode in diastolic heart failure model, Heart Fail. Rev. (2021), doi:10.1007/s10741-021-10117-6.

47. A. Escher, Y. Choi, F. Callaghan, B. Thamsen, U. Kertzscher, M. Schweiger, M. Hübler, M. Granegger, A Valveless Pulsatile Pump for Heart Failure with Preserved Ejection Fraction: Hemo- and Fluid Dynamic Feasibility, Ann. Biomed. Eng. (2020), doi:10.1007/s10439-020-02492-2.

48. F. Vincent, J. Ternacle, T. Denimal, M. Shen, B. Redfors, C. Delhaye, M. Simonato, N. Debry, B. Verdier, B. Shahim, T. Pamart, H. Spillemaeker, G. Schurtz, F. Pontana, V. H. Thourani, P. Pibarot, E. Van Belle, Transcatheter Aortic Valve Replacement in Bicuspid Aortic Valve Stenosis, Circulation 143, 1043–1061 (2021).

49. A. W. Harris, P. Pibarot, C. M. Otto, Aortic Stenosis: Guidelines and Evidence Gaps, Cardiol. Clin. 38, 55–63 (2020).

50. L. Rosalia, K. K. Lamberti, M. K. Landry, C. M. Leclerc, F. D. Shuler, N. C. Hanumara, E. T. Roche, A Soft Robotic Sleeve for Compression Therapy of the Lower Limb, Annu. Int. Conf. IEEE Eng. Med. Biol. Soc. IEEE Eng. Med. Biol. Soc. Annu. Int. Conf. 2021, 1280–1283 (2021).

