## Supplementary for "Tunable, soft robotics-enabled patient-specific hydrodynamic model of aortic stenosis and secondary ventricular remodeling"

### Supplementary materials

**Table S1. Summary of patients' structural and functional characteristics.** Left ventricular end-diastolic diameter (LVEDD), left ventricular end-systolic diameter (LVESD), and metrics of aortic stenosis (AS) were obtained from echocardiography imaging. Annular diameter, ascending aortic diameter, and left ventricular end-diastolic volume (LVEDV) were obtained from computed tomography imaging. Stroke volume (SV) was estimated from echocardiography imaging using the Teicholz equation and the left ventricular ejection fraction (LVEF) was calculated from values of SV and LVEDV. Left ventricular pressure (LVP) (peak/end-systolic/end-diastolic) and aortic pressure (AoP) (systolic/diastolic/mean) were obtained from catheterization data when available. NA: not available.

| **Patient** | **Structural parameters** | | | | | **Cardiac function** | | **AS metrics** | | | | **Cath data** | |
|---|---|---|---|---|---|---|---|---|---|---|---|---|---|
| | LVEDD (mm) | LVESD (mm) | Annulus diameter (mm) | Ascending aortic diameter (mm) | LVEDV (mL) | SV (mL) | LVEF (%) | EOA ($cm^2$) | $\Delta P_{mean}$ (mmHg) | $\Delta P_{max}$ (mmHg) | $v_{max}$ (cm/s) | LVP (mmHg) | AoP (mmHg) |
| 1 | 47 | 34 | 24 | 37.6 | 127 | 54.9 | 43.2 | 0.8 | 34 | 58 | 3.8 | NA | NA |
| 2 | 55 | 42 | 28 | 38.2 | 200 | 68.8 | 34.4 | 0.9 | 58 | 93 | 4.5 | NA | NA |
| 3 | 46 | 34 | 23 | 31.8 | 123 | 49.9 | 40.6 | 0.8 | 61 | 95 | 4.9 | NA | NA |
| 4 | 36 | 23 | 22 | 36.3 | 49 | 36.3 | 73.6 | 1.1 | 29 | 56 | 3.9 | NA | NA |
| 5 | 38 | 24 | 24 | 40.1 | 88 | 41.7 | 47.2 | 0.5 | 43 | 71 | 4.3 | 181/9/18 | 126/63/89 |
| 6 | 52 | 31 | 25 | 35.4 | 127 | 91.5 | 71.8 | 0.9 | 46 | 81 | 4.6 | NA | NA |
| 7 | 44 | 26 | 26 | 38.2 | 112 | 63.0 | 56.2 | 0.9 | 39 | 65 | 4.0 | NA | NA |
| 8 | 47 | 27 | 23 | 40.1 | 147 | 75.3 | 51.2 | 0.7 | 40 | 63 | 3.9 | NA | NA |
| 9 | 41 | 23 | 25 | 32.2 | 128 | 56.1 | 44.0 | 0.8 | 48 | 78 | 4.4 | NA | NA |
| 10 | 30 | 21 | 27 | 34.4 | 69 | 20.5 | 29.5 | 0.7 | 29 | 57 | 3.9 | NA | NA |
| 11 | 48 | 36 | 22 | 37.3 | 129 | 53.0 | 41.1 | 0.8 | 18 | 31 | 2.8 | 156/8/15 | 129/77/102 |
| 12 | 44 | 28 | 30 | 44.7 | 180 | 58.1 | 32.3 | 0.8 | 64 | 100 | 5.8 | 197/24/36 | 112/63/81 |
| 13 | 65 | 53 | 32 | 40.7 | 258 | 80.6 | 31.3 | 1.1 | 34 | 57 | 3.8 | 141/7/27 | 91/61/72 |

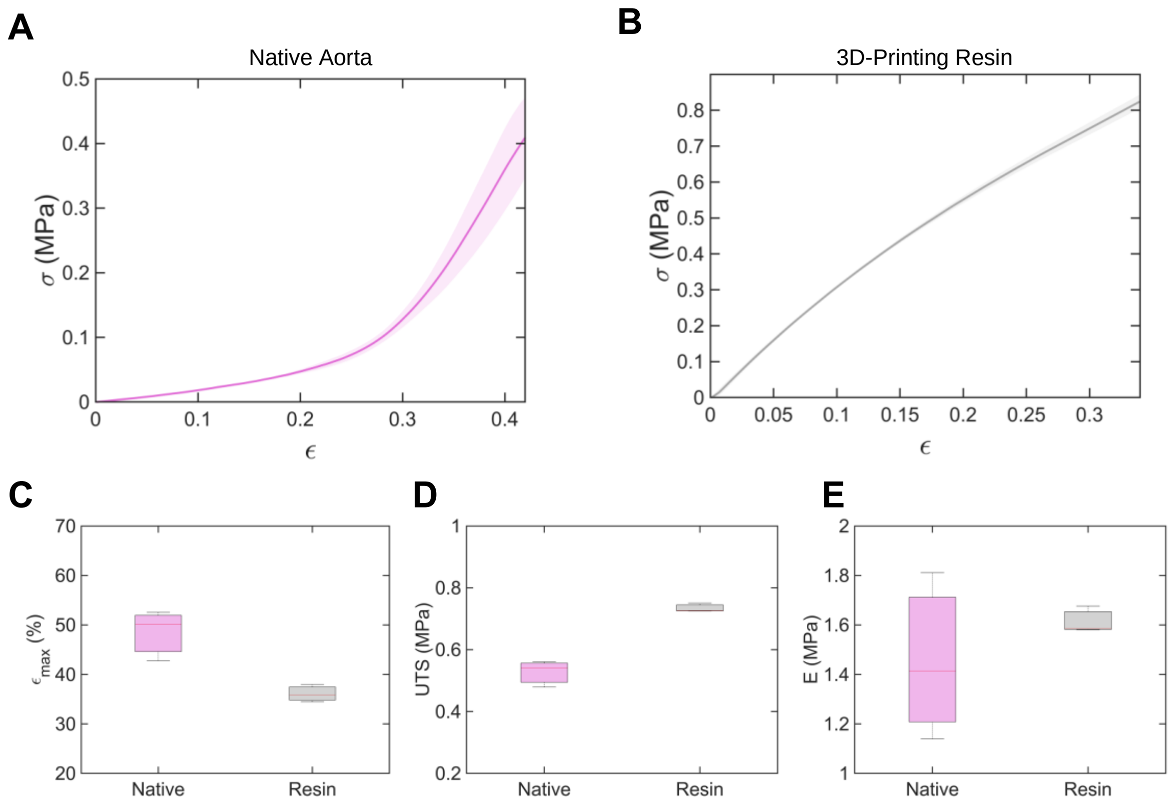


**Fig. S1. Mechanical characterization of the native porcine aorta and 3D-Printing resin**. Stress-strain raw data of (**A**) the native aorta and of the (**B**) the 3D-printing resin. Mechanical testing was performed using an electromechanical tester (Instron 5566, 50N load cell, Norwood, USA), and dogbone-shaped specimens subjected to uniaxial loading at a rate of 2 mm/min according to the American Society for Testing and Materials (ASTM) standards. Graphs show mean ± 1 s.d. (n = 3). **(C)** Maximum specimen elongation at failure, (**D**) Ultimate tensile strength (UTS), (**E**) Young's modulus for three different specimens of the native aorta and the resin (n = 3). Young's modulus was calculated in the 25-35% strain interval. σ: engineering stress, ε: engineering strain, E: Young's modulus.

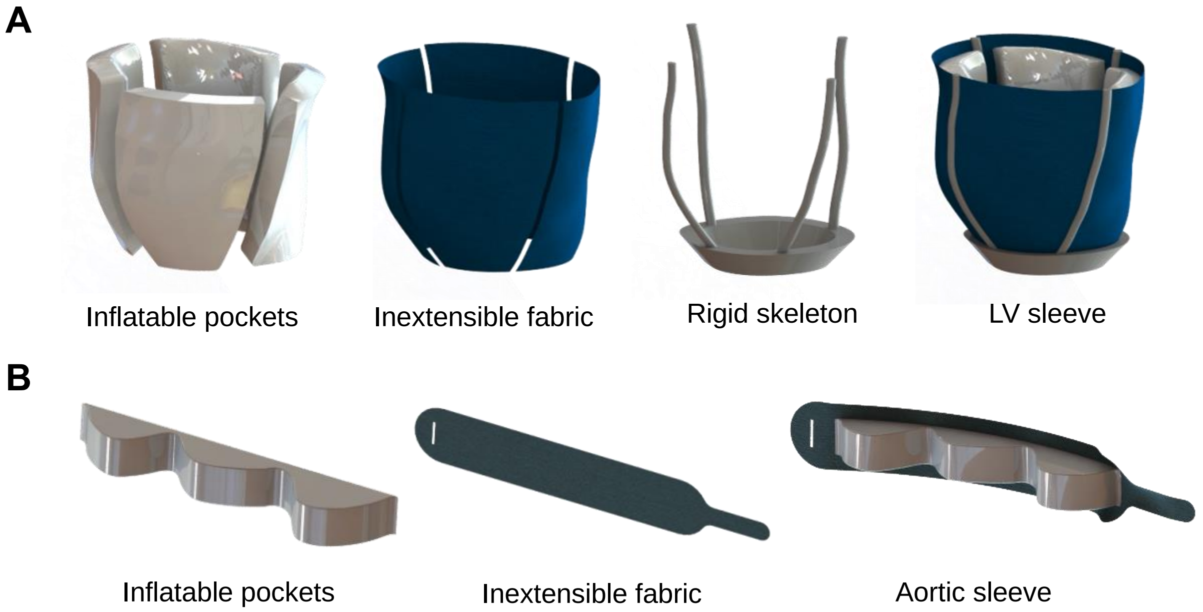


**Fig. S2. Design of LV and aortic sleeves**. (**A**) Inflatable TPU pockets, inextensible fabric layer, rigid skeleton, and assembly of the LV sleeve. (**B**) Inflatable TPU pockets, inextensible fabric, and assembly of the aortic sleeve.

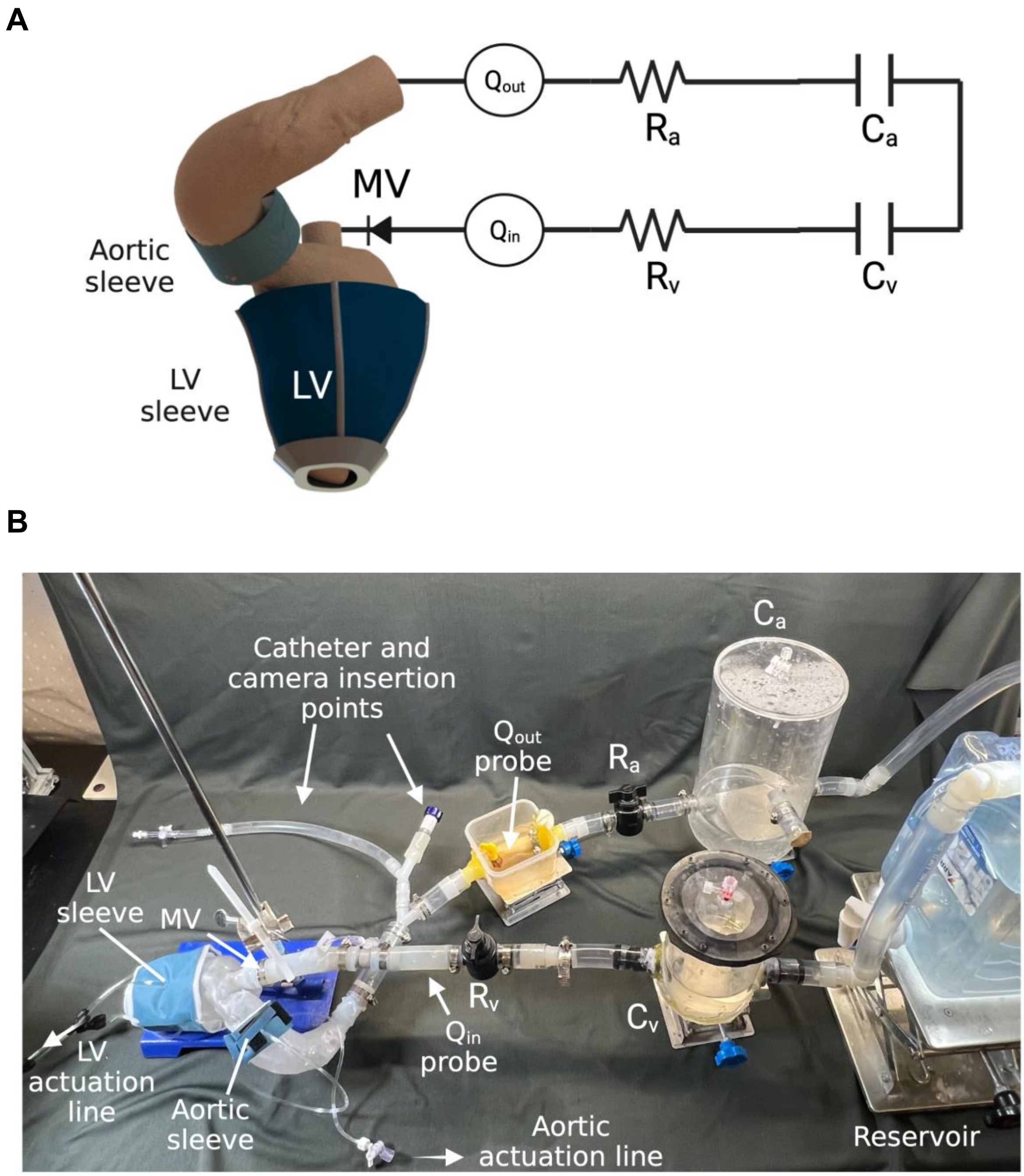


**Fig. S3. Hydrodynamic loop**. (**A**) Schematic of hydrodynamic loop illustrating the 3D-printed patient-specific anatomy of the left ventricle (LV) and aorta, the LV sleeve, the aortic sleeve, the outflow flowmeter ($Q_{out}$), the arterial resistance ($R_a$) and compliance ($C_a$), the venous resistance ($R_v$) and compliance ($C_v$), the inflow flowmeter ($Q_{in}$), and the mitral valve (MV). (**B**) Illustration of the physical hydrodynamic loop illustrating the components listed in (A), as well as the LV and aortic actuation lines, the catheter and camera insertion points, and the reservoir.

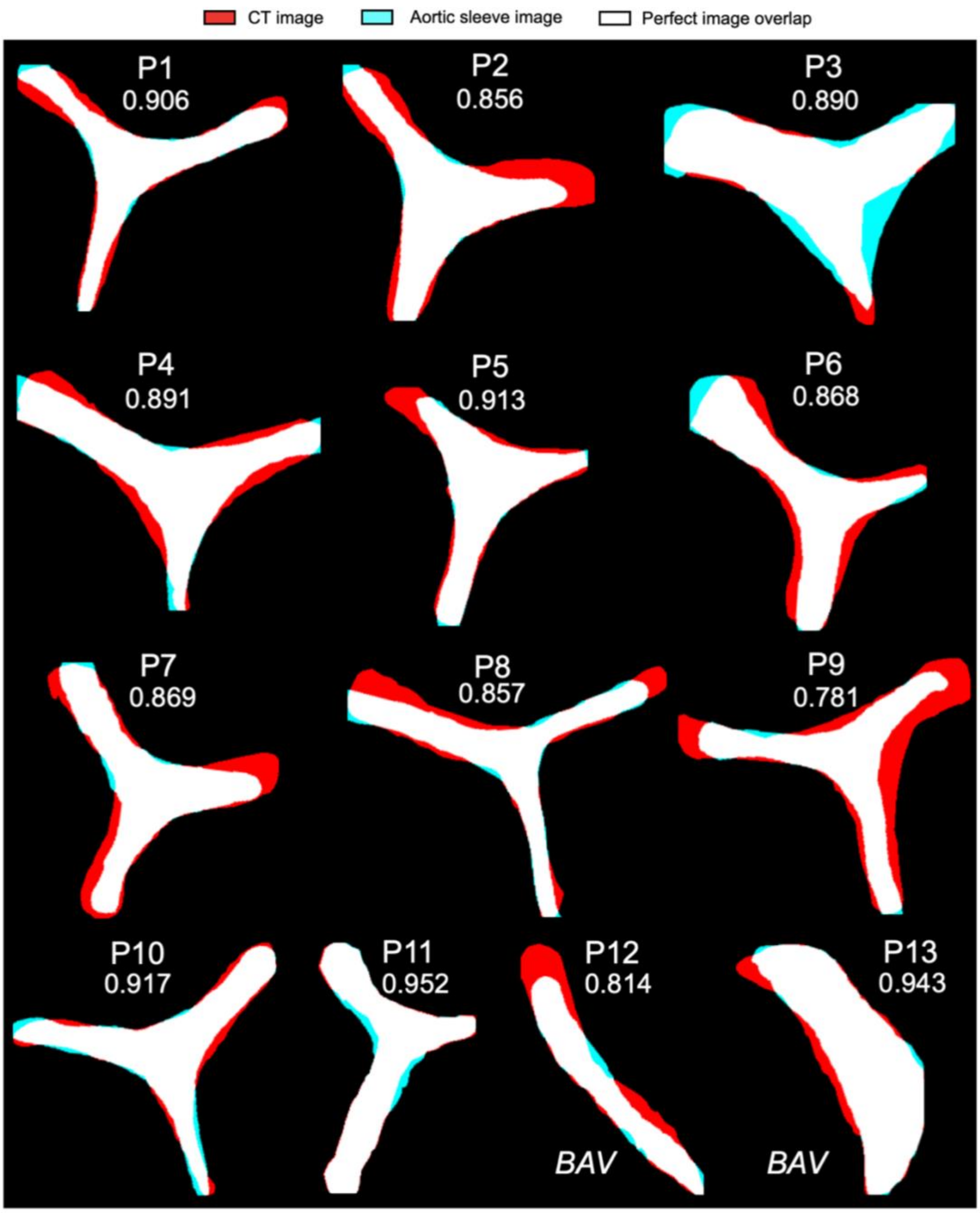


**Fig. S4. Comparison of aortic stenosis morphologies obtained by computed tomography (CT) and actuation of the soft robotic aortic sleeve**. Images for all the 13 patients (P 1-13) were binarized and cross-registered. White regions correspond to a perfect overlap between each pair of images, while cyan or red regions correspond to areas of noncongruent geometries. For each pair of images, the DSC was computed and was reported below the corresponding patient number. BAV: bicuspid aortic valve.

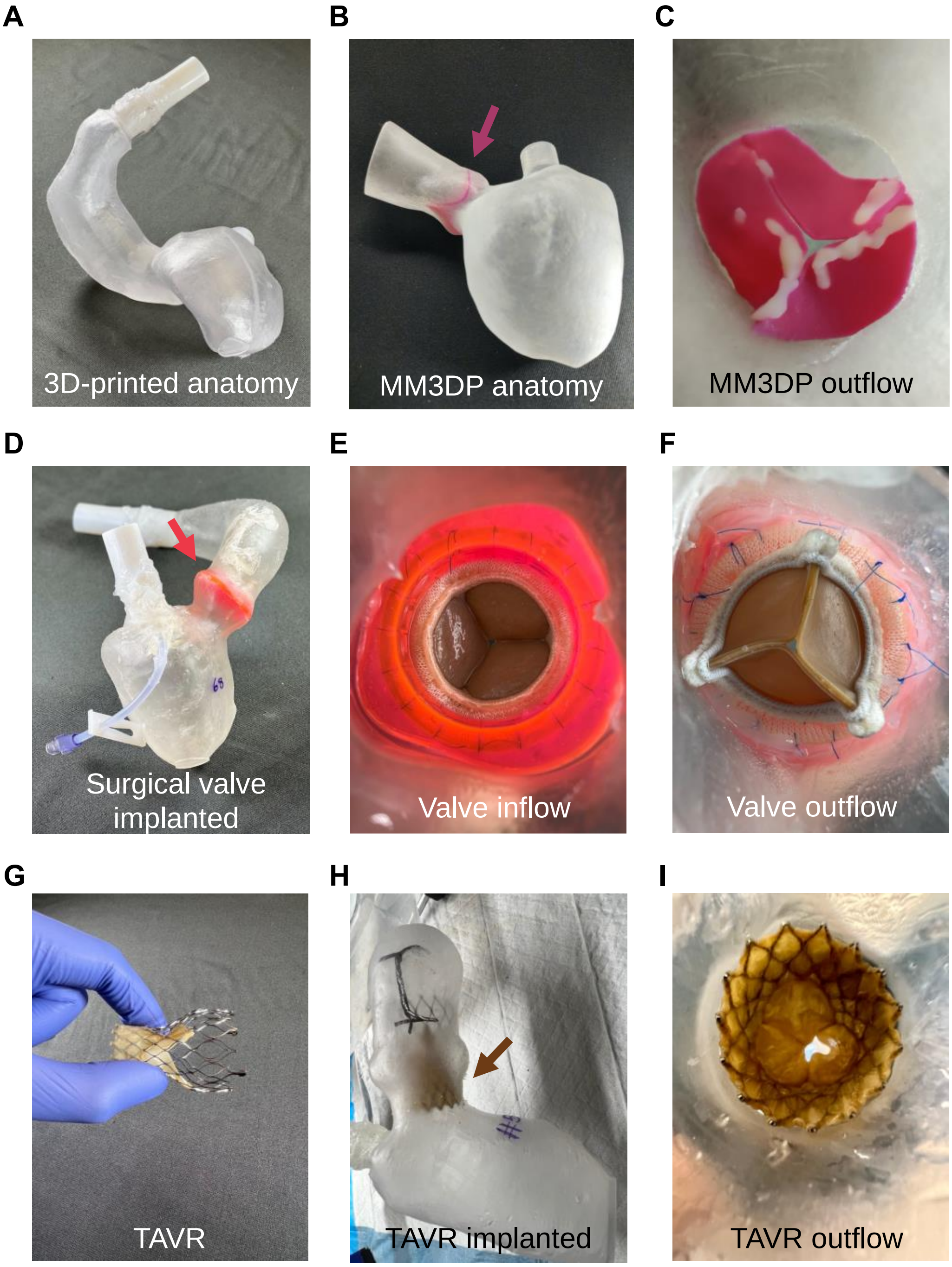


**Fig. S5. Illustration of 3D printing methods and use of surgical and TAVR valves**. (**A**) 3D-printed anatomy of the LV and aorta of use for soft robotics-driven model. (**B**) Multi-material 3D-printed (MM3DP) anatomy of the LV, aorta, and aortic valve (arrow). (**C**) Details of the MM3DP valve outflow. (**D**) Surgical valve implanted (arrow) in 3D-printed anatomy to recreate physiologic aortic hemodynamics, with details of the implanted valve (**E**) inflow and (**F**) outflow. (**G**) View of the Evolut R (Medtronic) self-expandable TAVR. (**H**) TAVR implanted (arrow) in the 3D-printed anatomy. (**I**) Detail of the TAVR in outflow view.

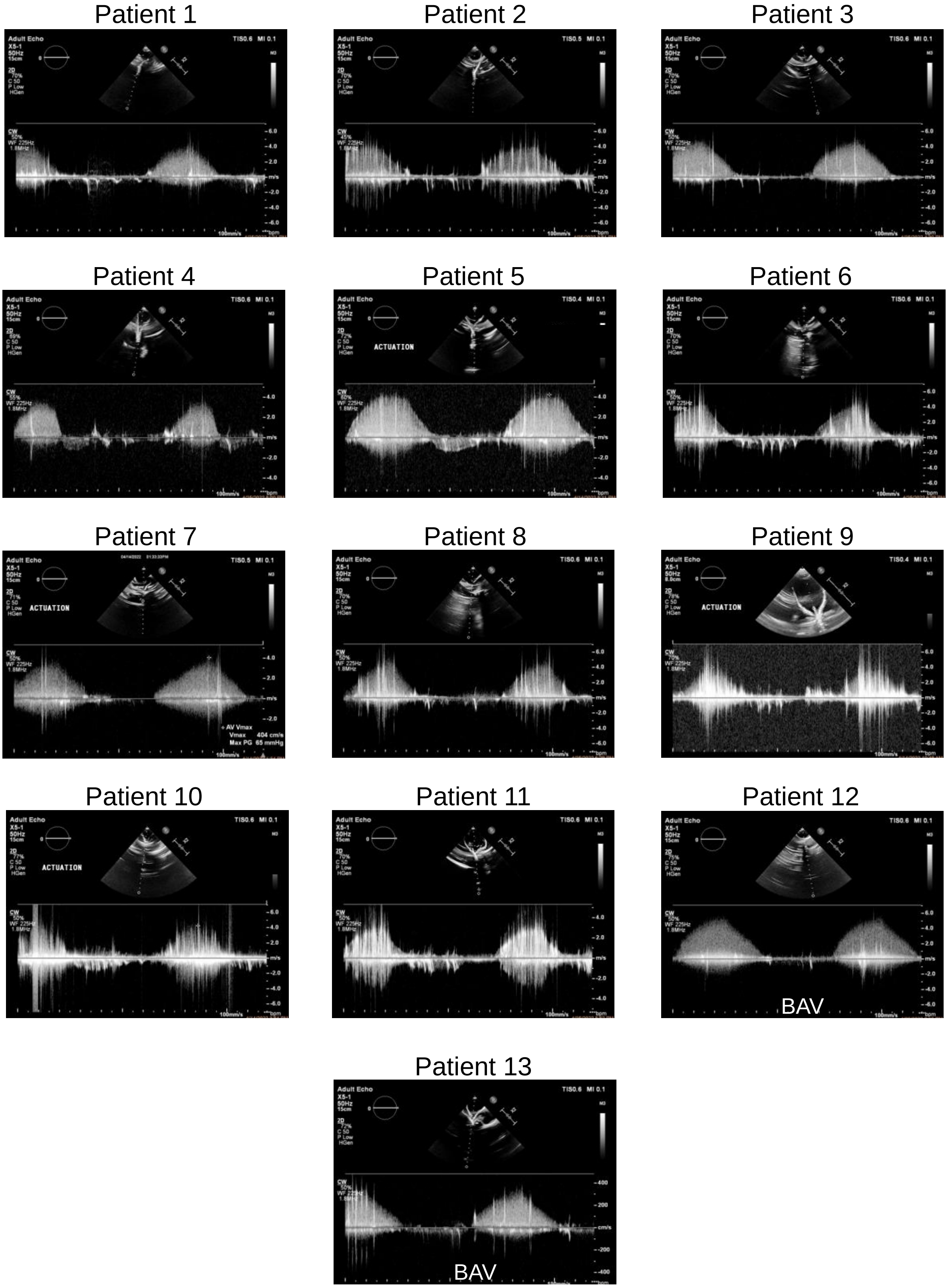


**Fig. S6. Representative echocardiographic data of soft robotics-driven hydrodynamic model**. Continuous-wave (CW) Doppler echocardiography on patients 1 – 13 for evaluation of peak blood flow velocity, as a critical metric for the evaluation of severity of aortic stenosis. BAV: bicuspid aortic valve.
